# The Effectiveness and Cost-Effectiveness of Community Health Workers in Delivering Antenatal Care and Immunization Services to Pregnant Mothers and Children Under 5 in Rural Africa: A Study Protocol for a Systematic Review and Meta-Analysis

**DOI:** 10.1101/2024.08.18.24312186

**Authors:** Kebby Zumani, Henry Irumba, Adebukunola Olajumoke Afolabi, Semkelokuhle Dube, Vincent Byaruhanga, Olaotuyombo George, Mark Anum Nortey, Emmanuel Owusu Ofori, Birungi Bk Valentine, Patience Peru, Chikosa Kamwengo, Amoah Roberta Mensima

## Abstract

**Background:** Maternal and child health care, particularly antenatal care (ANC) and immunization services, are essential to improving health outcomes in rural Africa. Despite global efforts, access to high-quality health care services remains limited in rural areas, contributing to high maternal and childhood mortality rates. Community Health Workers (CHWs) have been recognized as a promising solution for bridging this gap by providing essential services directly to underserved populations. This systematic review and meta-analysis aims to evaluate the effectiveness and cost-effectiveness of CHWs in delivering antenatal care and immunization services to pregnant mothers and children under 5 in rural Africa.

**Methods:** This review will include randomized controlled trials (RCTs), cohort studies, case-control studies, and observational studies published from 2014 onward. The search strategy will be implemented across multiple databases, including Google Scholar, Academic Info, Cochrane, Refseek, PubMed, and MEDLINE. The primary outcomes will focus on clinical and economic measures, including maternal and child health outcomes and cost-effectiveness of CHW interventions. Data extraction and quality assessment will be conducted independently by two reviewers, with discrepancies resolved through discussion or the involvement of a third reviewer.

**Discussion:** The findings from this review will contribute to the understanding of the role CHWs play in improving maternal and child health outcomes in rural Africa. The results will provide valuable insights for policymakers, health care providers, and stakeholders to inform future interventions and resource allocation strategies.

**Registration:** This protocol is registered in PROSPERO Registration number CRD42024529963.

## 1.0. Introduction

Antenatal care and immunization are essential health care services for pregnant women and children. These services play a crucial role in promoting health, diagnosing and preventing diseases, and improving health outcomes. According to the World Health Organization (WHO) and the United Nations Children’s Fund (UNICEF), every pregnant women deserves higher quality ANC services, and all children need immunization to protect them from infectious diseases. Unfortunately, maternal and childhood remains a significant concern globally, with more than two thirds of maternal deaths occurring in Africa (WHO, 2023). Rural African communities face several challenges, including limited health care infrastructure, socioeconomic disparities and geographical remoteness, which make accessing qualitative care difficult (Adekdokun et al, 2023 : sahoo et al, 2021). As such, improving maternal and child health remains a global public health agenda for the sustainable development goals (Akalewold et al, 2022).

Community health workers (CHWs) have emerged as a critical resource in bridging the gap in healthcare accessibility, especially in rural regions. Often selected from within communities they serve, CHWs play a vital role in delivering essential services directly to beneficiaries. The WHO defines community health workers as front-line public health workers who have a deep understanding of the community they serve and are trusted members of that community. Community health workers serve as a link between health and social services and community to facilitate access to services and improve the quality and the cultural competencies of service delivery (peprah et al, 2020). In the context of maternal and child health, community health workers are increasingly being utilized to provide the antonym of care and immunization services, addressing the unique challenges posed by the geographical and social economic landscape of rural Africa (Roux et al, 2020).

Integrating community health workers into the Healthcare System has shown promise in enhancing the coverage and effectiveness of antenatal care and immunisation services. Their deep rooted connection to local communities enable them to facilitate trust, culturally sensitive communications, and the delivery of healthcare services tailored to the specific needs of pregnant women and young children (Olaniran et al, 2019 : WHO, 2023). Moreover, CHWs Potential cost -effectiveness in comparison to traditional healthcare and models makes their role even more compelling, particularly in resource-limited settings (Perry et al, 2021). Studies have shown that community health workers are a less expensive alternative to other cadres of health workers, with lower salary and training costs (Van Ginneken et al., 2013).

However, despite the growing recognition of CHWs as valuable assets in maternal and child healthcare, a comprehensive synthesis of existing evidence through a systematic review and meta-analysis is necessary (Olaniran et al., 2019; WHO, 2023) to understand the effectiveness and cost-effectiveness of CHWs in delivering these services fully. This understanding will help fill existing gaps in the literature, inform policy and practice and guide future interventions aimed at improving maternal and child health outcomes in rural communities

This systematic review and meta-analysis aims to synthesize the available evidence on the effectiveness and cost-effectiveness of CHWs in delivering antenatal care and immunization services in rural Africa. The findings will guide future policy and practice, addressing the need for improved maternal and child health outcomes in these underserved regions.

## 2.0. Research Question and Objectives

### 2.1. Research Question

What is the effectiveness and cost-effectiveness of CHWs in delivering antenatal care and immunization services to pregnant mothers and children under 5 in rural Africa, and what factors influence the outcomes of these interventions?

### 2.2. Study Objectives

- To evaluate the effectiveness of community health worker interventions on antenatal care and immunization outcomes among pregnant mothers and children under 5 in rural Africa.
- To evaluate the cost-effectiveness of community health worker interventions on antenatal care and immunization outcomes among pregnant mothers and children under 5 in rural Africa.
- To assess the associated factors influencing the effective and cost effective delivery of antenatal care and immunization services by community health workers to pregnant women and children under 5 in rural Africa

## 3.0. Methods

### 3.1. Eligibility Criteria

#### Inclusion Criteria

- **Study Types:** Randomized controlled trials (RCTs), cohort studies, case-control studies, and observational studies.
- **Population:** Pregnant mothers and children under 5 years in rural Africa.
- **Intervention:** CHWs delivering antenatal care and immunization services.
- **Comparators:** Traditional health care models, alternative community-based interventions, or standard care without CHW involvement.
- **Outcomes:** Clinical outcomes (maternal and child health outcomes) and economic outcomes (cost-effectiveness, resource utilization).
- **Publication Date:** Studies published from 2014 onward.

#### Exclusion Criteria

- **Study Types:** Reviews, editorials, studies lacking primary data, and studies with insufficient information for data extraction and analysis.
- **Population:** Studies conducted in urban or non-African settings, or focusing on populations outside the specified age groups.
- **Intervention:** Studies that do not specifically evaluate CHW interventions related to antenatal care and immunization services.
- **Outcomes:** Studies lacking relevant outcome measures or without clear reporting on the effectiveness or cost-effectiveness of CHW interventions.

### 3.2. Search Strategy

The search will be conducted across multiple databases, including Google Scholar, Academic Info, Cochrane, Refseek, PubMed, and MEDLINE. Search terms will include combinations of the following keywords: “Community health workers,” “Antenatal care,” “Immunization,” “Rural Africa,” “Village health workers,” and “Sub-Saharan Africa.” A multistage search approach will be employed, with a focus on identifying studies published within the last 10 years.

### 3.3 Data Extraction

Two reviewers will independently extract data using a standardized form. A cross-verification process will be implemented to address any discrepancies. Quality assessments will be conducted independently, with disagreements resolved through discussion or the involvement of a third reviewer.

### 3.3. Risk of Bias Assessment

Quality assessment will be guided by the Joanna Briggs Institute (JBI) Critical Appraisal Tools, evaluating study design, sample size, representativeness, bias and confounding, data collection methods, and outcome measures.

### 3.4. Data Synthesis

A narrative synthesis will be conducted to group similar studies and summarize key findings. Meta-analysis will be performed where appropriate, using statistical methods to combine results from comparable studies. Sensitivity analysis will be conducted to assess the robustness of the findings, particularly in studies with missing or unclear data.

## 4.0. Discussion

This systematic review and meta-analysis will provide a comprehensive understanding of the role of CHWs in delivering antenatal care and immunization services in rural Africa. The findings will contribute to the evidence base on the effectiveness and cost-effectiveness of CHW interventions, offering valuable insights for policymakers, health care providers, and stakeholders. The results will guide future interventions aimed at improving maternal and child health outcomes in underserved regions, aligning with global public health agendas, including the Sustainable Development Goals (SDGs).

## 5.0. Ethical Considerations

For developing this review, formal ethics approval is not required as the analysis would be based on reviewing existing literature on published articles related to the effectiveness and cost effectiveness of community health workers in delivering ANC and immunization services to pregnant women and children under 5 in rural Africa. The study does not involve collection of personal, sensitive or confidential information from participants henceforth written informed consent is not required.

The authors declare that appropriate inclusion and exclusion criteria would be used in selection of studies related to the review title. Through purposefully informed selective inclusivity, reviewers shall distil information that is most relevant for addressing the research questions to minimize search and publication bias.

Reviewers shall ethically evaluate the extent to which findings reported in individual studies are grounded in the reported evidence. Transparency and honesty shall be observed in communicating the insights gained from this study. Any conflicts of interest that may arise during the review process shall be communicated and resolved. Authors shall acknowledge all forms of external contributions to the research.

## 6.0. Dissemination Plan

The findings of this systematic review and meta-analysis will be disseminated through multiple channels to ensure that they reach a broad audience, including policymakers, healthcare providers, community health workers (CHWs), researchers, and the general public. The dissemination plan includes the following strategies:

Publication in Peer-Reviewed Journals: The final study results will be submitted for publication in reputable peer-reviewed journals specializing in public health, maternal and child health, and health economics. Target journals may include those with a focus on global health, healthcare interventions in low-resource settings, and systematic reviews.

Open access publication will be prioritized to ensure that the findings are accessible to a wider audience, particularly in low- and middle-income countries.

Conference Presentations: The research team will present the findings at national and international conferences related to public health, maternal and child health, and global health. These presentations will facilitate the sharing of knowledge and foster discussions among experts and stakeholders in the field.

Workshops and Seminars: The research team will organize workshops and seminars in collaboration with academic institutions, governmental organizations, and non-governmental organizations (NGOs) across Africa. These sessions will target local healthcare providers, CHWs, and policymakers to discuss the implications of the findings and explore strategies for implementing evidence-based practices.

Policy Briefs and Reports: A series of policy briefs and reports summarizing the key findings and recommendations will be developed. These documents will be tailored for policymakers and healthcare administrators in rural African regions, highlighting actionable steps to enhance the effectiveness and cost-effectiveness of CHW interventions in antenatal care and immunization services.

The reports will also be shared with international organizations such as WHO, UNICEF, and local health ministries.

Digital and Social Media: The research findings will be disseminated through digital platforms, including the Afromed Academy of Medical Research and Innovation’s website and social media channels. Infographics, blog posts, and short videos will be created to communicate the key results in a more engaging and accessible manner.

Academic networking sites such as ResearchGate and LinkedIn will also be used to share the research outcomes with the global research community.

Engagement with Funders and Stakeholders: The findings will be shared with potential funders, donors, and stakeholders who are interested in supporting initiatives that improve maternal and child health outcomes in rural Africa. Presentations and meetings will be organized to discuss the potential for scaling up successful CHW interventions based on the study’s results.

Contribution to Databases and Repositories: The data and findings will be made available through relevant academic and public health databases, such as PROSPERO, the Cochrane Library, and other systematic review repositories. This will ensure that the research is accessible to other researchers and contributes to the broader body of evidence in the field.

This multifaceted dissemination plan aims to maximize the impact of the research, ensuring that the findings inform policy, practice, and future research in maternal and child health across rural Africa and beyond.

## Data Availability

All data produced in the present study are available upon reasonable request to the authors

https://www.crd.york.ac.uk/PROSPERO/display_record.php?RecordID=529963

## Acknowledgments

We acknowledge the contributions of all team members and affiliated institutions for their dedication to this project. Special thanks also goes to all members of the academic faculty at Afromed Academy of Medical Research and Innovation.

## References

1. Adedokun, S. T., Adekanmbi, V. T., Uthman, O. A., Lilford, R. J., & Adetokunboh, O. O. (2023). Spatial distribution of antenatal care utilization and maternal health outcomes in sub-Saharan Africa: A systematic review and meta-analysis. BMC pregnancy and childbirth, 23(1), 1–15.

2. Corluka, A., Walker, D. G., Lewin, S., Glenton, C., & Scheel, I. B. (2009). Are vaccination programmes delivered by lay health workers cost-effective? A systematic review. Human resources for health, 7, 81. 10.1186/1478-4491-7-81

3. Kaiser, J., & Barstow, D. (2022). The pandemic’s frontline heroes. Science, 371(6532), 22–27.

4. le Roux, K. W., Mbatha, J. N., & Ntuli, S. A. (2020). Community health workers’ experiences of using video-based mobile learning to improve maternal and child health services in rural KwaZulu-Natal, South Africa. African Journal of Primary Health Care & Family Medicine, 12(1), 1–7.

5. Muheirwe, F. & Nuhu, A. (2019). Antenatal care services utilization and factors affecting it among mothers in rural communities of Uganda. Journal of Public Health in Africa, 10(1), 1–8.

6. Olaniran, A., Smith, H., Unkels, R., Bar-Zeev, S., & van den Broek, N. (2019). Who is a community health worker?–a systematic review of definitions. Global health action, 12(1), 1–12.

7. Pegurri, E., Fox-Rushby, J. A., & Damian, W. (2005). The effects and costs of expanding the coverage of immunisation services in developing countries: a systematic literature review. Vaccine, 23(13), 1624–1635. 10.1016/j.vaccine.2004.02.029

8. Perry, H. B., Sacks, E., Schleiff, M., Kumapley, R., Gupta, S., Rassekh, B. M., & Freeman, P. A. (2021). Comprehensive review of the evidence regarding the effectiveness of community–based primary health care in improving maternal, neonatal and child health: 6. strategies used by effective projects. Journal of global health, 11(1), 1–16.

9. Sahoo, K. C., Sahoo, S., Choudhury, A. K., Sofi, N. Y., Kumar, R., & Bhattacharya, S. K. (2021). Childhood immunization in Odisha, India: a systematic review and meta-analysis. BMC public health, 21(1), 1–14.

10. Tseng, Y. H., Griffiths, F., de Kadt, J., & Hill, P. C. (2019). Understanding the role of community health workers in improving maternal and child health in low-income settings: protocol for a systematic review and qualitative synthesis. Systematic reviews, 8(1), 1–8.

11. UNICEF. (2022). Antenatal care. Retrieved from https://www.unicef.org/early-childhood-development/antenatal-care

12. Van Ginneken, N., Lewin, S., Berridge, V., & Tumwijukye, I. (2013). The emergence of community health worker programmes in the late apartheid era in South Africa: An historical analysis. Social Science & Medicine, 97, 250–258.

13. WHO. (2023). Maternal health. Retrieved from https://www.who.int/health-topics/maternal-health.

